# Indirect effects of the COVID-19 pandemic on paediatric health-care use and severe disease: a retrospective national cohort study

**DOI:** 10.1101/2020.10.15.20212308

**Authors:** Thomas C Williams, Clare MacRae, Olivia Swann, Haris Haseeb, Steve Cunningham, Philip Davies, Neil Gibson, Christopher Lamb, Richard Levin, Catherine McDougall, Jillian McFadzean, Ian Piper, Alastair Turner, Steve Turner, Margrethe Van Dijke, Don Urquhart, Bruce Guthrie, Ross J Langley

**Author notes:** Contributed equally (Bruce Guthrie, Ross J Langley). **Corresponding author:** Ross J Langley, Department of Paediatric Respiratory and Sleep Medicine, Royal Hospital for Children, Glasgow.

## Abstract

**Background:** Severe disease directly associated with SARS-CoV-2 infection in children is rare. However, the indirect consequences of the COVID-19 pandemic on paediatric health have not been fully quantified. We examined paediatric health-care utilisation, incidence of severe disease, and mortality during the lockdown period in Scotland.

**Methods:** This national retrospective cohort study examined national data for emergency childhood primary and secondary care utilisation following national lockdown on March 23, 2020. To determine whether social distancing measures and caregiver behavioural changes were associated with delayed care-seeking and increased disease severity on presentation, unplanned, emergency admissions requiring invasive mechanical ventilation for the two national Paediatric Intensive Care Units (PICUs) were analysed. PICU admissions were grouped by diagnostic category, and disease severity on presentation calculated. National statutory death records were consulted to establish childhood mortality rates and causes of death. For all observations, the lockdown period was compared to equivalent dates in 2016-2019.

**Findings:** We identified 273,455 unscheduled primary care attendances; 462,437 emergency department attendances; 54,076 emergency hospital admissions; 413 PICU emergency admissions; and 415 deaths during the lockdown study period and equivalent dates in previous years. The rates of emergency presentations to primary and secondary care fell during lockdown in comparison to previous years. Emergency PICU admissions for children requiring invasive mechanical ventilation also fell, with an odds ratio of 0·52 for chance of admission during lockdown (95% CI 0·37-0·73, p < 0·001). Clinical severity scores did not suggest children were presenting with more advanced disease. The greatest reduction in PICU admissions was for diseases of the respiratory system; those for injury, poisoning or other external causes were equivalent to previous years. Mortality during lockdown did not change significantly compared to 2016-2019.

**Interpretation:** National lockdown led a reduction in paediatric emergency care utilisation, without associated evidence of severe harm.

**Funding:** None

**Research in context:** *Evidence before this study:* Data on the indirect effects of the COVID-19 pandemic on children at a population level are limited. We searched PubMed and medRxiv on October 13, 2020, for studies published from Jan 1, 2020 examining the indirect effects of non-pharmaceutical interventions (NPIs), and associated changes in caregiver health-care seeking behaviour, on the risk of severe paediatric disease and death. We used the search terms COVID-19, SARS-CoV-2, non-pharmaceutical interventions, indirect, and children, as well as manually searching references in other relevant papers. Terms were searched individually and in combination as necessary, with no language restrictions. We identified one study that modelled the indirect effects of the COVID-19 pandemic on child deaths in low- and middle-income countries. Other studies analysed in isolation the effects of NPIs and other behavioural changes on emergency department attendances, hospital admission rates, paediatric intensive care unit (PICU) admission rates, or the incidence of specific presentations, such as asthma exacerbations. Some case series described delayed care-seeking for children with non-SARS-CoV-2 disease. We did not identify any national studies examining the indirect effects of the COVID-19 pandemic on the incidence of severe paediatric disease and mortality.

*Added value of this study:* This national study quantified the changes following national lockdown in Scotland on March 23, 2020. We examined data for unscheduled primary care and emergency department attendances, emergency hospital admissions, emergency paediatric intensive care unit (PICU) admissions requiring invasive mechanical ventilation, and paediatric mortality. Rates were compared with previous years. We found a reduction in paediatric emergency care utilisation rates associated with national lockdown. This reduction is likely to be due to a combination of changes in health care seeking behavior, and a fall in overall burden of paediatric infectious disease. These measures did not appear to have been associated with evidence of severe harm to children in Scotland, as evidenced by severity scores on presentation to PICU or mortality.

*Implications of all the available evidence:* This is the first comprehensive population-based assessment at a national level of the indirect effects of the COVID-19 pandemic on severe paediatric morbidity and mortality. Despite a significant reduction in health-care utilisation rates, we did not find associated evidence of severe harm. This study will assist policy makers, health-care providers and the public in evaluating the effects of lockdown on the risk of severe paediatric disease at a population level.

## Introduction

The global impact of the coronavirus (SARS-CoV-2) pandemic (COVID-19) has been extensive with over 30 million confirmed cases and 1 million deaths globally as of October 2020.^1^ The first case of SARS-CoV-2 in Scotland was identified on March 1, 2020,^2^ with evidence of sustained community transmission ten days later.^3^ Sequencing data suggests the virus was introduced into Scotland from multiple European countries.^3^ With a rising number of cases across the United Kingdom, a UK-wide lockdown was implemented on March 23, 2020 (Figure 1A). This included a number of non-pharmacological interventions (NPIs), such as widespread closures of school/nurseries, leisure centres, indoor play spaces and non-essential workplaces, as well as promotion of home working, hand washing and wearing of face coverings ^4,5^ Phased easing of lockdown measures by the Scottish Government began on May 29, 2020; pupils returned to schools on August 12, 2020 (Figure 1B).^4^

**Figure 1.**
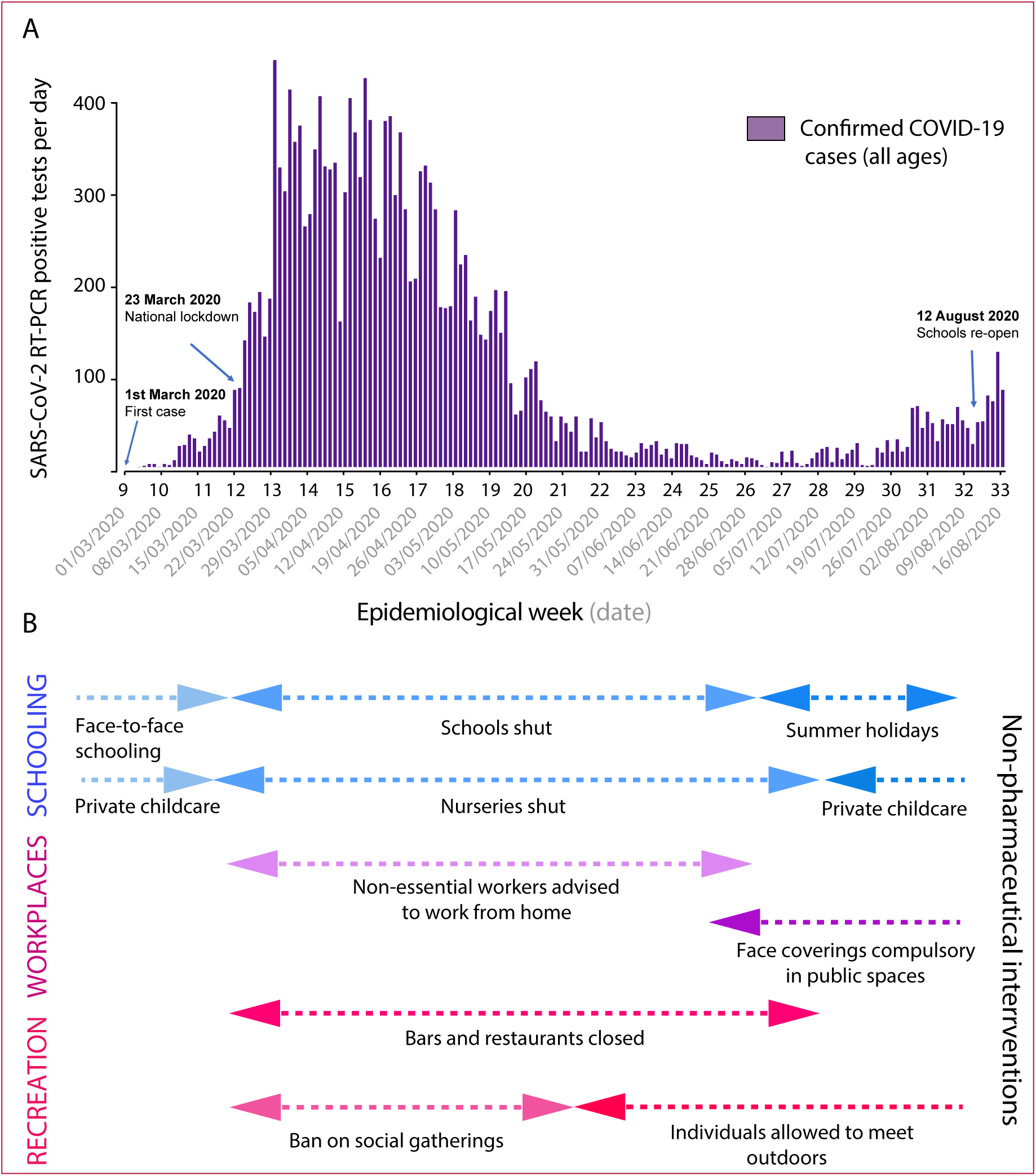
Annotated SARS-CoV-2 epidemic curve for Scotland (A) and key events during the lockdown period (B). (A) New positive SARS-CoV-2RT-PCR cases by day, annotated with key dates and events of relevance for this study. Data from Scottish Government.^30^ (B) Date ranges for key social distancing measures in Scotland during the lockdown period.^4^

Despite a significant direct impact of SARS-CoV-2 on adult mortality and morbidity, the paediatric population appeared to be relatively protected during the first wave of the pandemic, with a low number of reported infections, hospitalization rates and mortality.^6^ In England, which borders Scotland but has a population 10 times larger, there have been a small number (58) of reported cases of the pediatric inflammatory multisystem syndrome temporally associated with SARS-CoV-2 (PIMS-TS).^7^ Public Health Scotland (PHS) and National Records of Scotland (NRS) data shows that by the end of August 2020 in the 0-14 age range there had been 345 confirmed SARS-CoV-2 positive cases, 43 hospital admissions,^8^ and no deaths.^9^

Whilst severe disease directly associated with acute SARS-CoV-2 infection in children is rare,^6^ the health consequences of NPIs, widespread societal lockdown, and possible changes in caregiver health-care seeking behavior on children have not been fully quantified. A reduction in asthma exacerbations^10^ and PICU admissions for respiratory conditions have been seen,^11^ with both presentations commonly being triggered by respiratory viruses. Paediatric Emergency Department (ED) attendance rates in the Republic were reduced during the lockdown period compared to previous years.^12^ Other case series have shown delays in care-seeking behaviour^13^ and an increase in non-accidental injuries during lockdown,^14^ suggesting that there may be risks associated with decreased availability of health-care and lower levels of surveillance from health, education and social care workers during this period. Our study used complete national datasets to quantify the indirect effects of the COVID-19 pandemic on emergency paediatric health care utilisation rates and paediatric morbidity and mortality in Scotland during the lockdown period.

## Methods

### Notes on population

Scotland has an estimated population of 5,463,300.^15^ 96% of Scottish residents describe their ethnicity as “White”, and 4% describe themselves as being part of a minority ethnic group.^16^ 83% of the population are located in urban areas, mainly in the two main cities of Glasgow and Edinburgh.^16^

### Data sources and data collection

The overall design was a retrospective, population-based analysis of all emergency paediatric healthcare utilisation. We examined unscheduled primary care and emergency department attendances, emergency hospital admissions, emergency paediatric intensive care (PICU) admissions requiring invasive mechanical ventilation, and paediatric mortality for all children in Scotland. Disclosure controlled, aggregate count data of relevant healthcare utilisation events (apart from PICU admissions) was provided by Public Health Scotland (PHS). The data used is publicly available upon request, with no ethical permissions required. Emergency hospital admissions data are derived from the Rapid Preliminary Inpatient Data (RAPID) dataset. This dataset is submitted daily to PHS by NHS Boards across Scotland and provides data on patients admitted to general acute inpatient care, including under paediatric specialties. Unscheduled primary care, emergency department attendances and admissions to neonatal units are excluded from the RAPID dataset.^17^

Data from both PICUs in Scotland was analysed, and therefore our study provides complete national data for children requiring critical care. Information on unplanned, emergency admissions requiring invasive mechanical ventilation at the Royal Hospital for Children in NHS Greater Glasgow and Clyde and the Royal Hospital for Children and Young People in NHS Lothian, Edinburgh was collected. Analysis was performed as part of a service evaluation program and in concordance with both NHS Greater Glasgow and Clyde and NHS Lothian Quality Improvement policies ethical approval is not required. PICU data was collected in an electronic clinical information system (CIS) MetaVision® provided by iMDsoft® and in the Paediatric Intensive Care Audit Network (PICANet) database.^18^ National Records of Scotland provided detailed childhood mortality data, stratified by age group (0-4 and 5-14 years), and by cause of death classified by ICD-10 code. A number of deaths for 2019 and 2020 had not yet been reached final coding stage and therefore were documented as a “Not yet classified” group. National data on mid-year population estimates by age group were referenced from a National Records of Scotland (NRS) dataset.^19^

### Study design

#### Calculation of population at risk

The paediatric population was calculated by taking the number of children within each age category from the mid-year population estimates from NRS for each year. For 2020, the population at risk was estimated by taking the mid-year population estimates for years 0-13 for 2019, equating to ages 1-14 in 2020. To estimate the mid-year population of infants aged <1 in 2020, the ratio of the number of births in weeks 1-12 in 2020 to those in the equivalent period in 2019 was calculated and applied to the mid-year population for age group <1 year in 2019.

#### Study outcomes

Data for healthcare utilisation and mortality rates were examined for all children living in Scotland aged 0-14 years of age. Rates of health care use, including unscheduled primary care and emergency department attendance, and emergency hospital admissions were analysed. Healthcare utilisation rates from the period March 23 to August 9, 2020 (epidemiological weeks 13-32, corresponding to the start of lockdown, and the re-opening of schools) were compared to equivalent periods in previous years, calculated as a rate per 100,000 children. Unscheduled primary care (including telephone consultations) and emergency department attendances were available from January 1, 2016 onwards. Emergency hospital admission rates were obtained from the RAPID dataset for years 2018-2020. Attendance and admission rates in all settings were calculated by referencing the same epidemiological weeks for previous years.

Data referencing diagnosis codes for PICU admissions were available until June 30, 2020. The rate of admissions for the period March 23 to June 30, 2020 was compared to the equivalent epidemiological weeks of 2016-2019. Admissions diagnoses were classified according to ICD-11 diagnostic codes.^20^ Illness severity at presentation to PICU was determined by reference to the Paediatric Index of Mortality 2 (PIM2R) Score,^21^ calculated for purposes of submission to PICANet using a recalibration dating to 2016 (PIM2R 2016).^22^ As PICU admissions occur up to the age of 16 years, the population at risk used for calculations was that of children aged 0-15 years. Childhood mortality data, categorised by age groups 0-4 and 5-14 years, was available for epidemiological weeks 13-30 in 2020 (March 23 to July 31) and the equivalent periods in 2016-2019 for comparison. Codes for the cause of death were categorised by ICD-10 code.^23^ Mortality rates were normalised by the total population at risk as described above.

#### Statistical analysis

Statistical analysis was conducted using R,^24^ v3.4.1 and Graphpad Prism 6.04 (La Jolla California). For unscheduled primary care consultations, ED attendances and emergency admissions, geometric means with 95% confidence intervals were used to compare epidemiological weeks 5-32 (February 2 to August 9) of 2020 with equivalent weeks in 2016-2019. Two-sided Fisher’s exact tests with an alpha value of 0.05 were used to compare events in the lockdown period to previous years (epidemiological weeks 13-32, equating to March 23 to August 9, 2020). PICU admissions were analysed using geometric means with 95% confidence intervals (CIs) used to compare admissions by month (February to June) of 2020 to equivalent months in 2016-2019. A Fisher’s exact test was used to compare the number of admissions from March 23 to June 30, 2020, compared to admissions during equivalent weeks in 2016-2019. A non-parametric Kolmogorov-Smirnov test was used to compare mean severity score on presentation to PICU for these admissions for the same periods. Childhood mortality rates from epidemiological weeks 13-30 in 2020 (March 23 to July 26) were compared to the equivalent periods in 2016-2019, including calculation of 95% CIs. A z-score was calculated for childhood deaths and PICU admissions by category in 2020, by subtracting the 2016-2020 mean from the value for 2020 and by dividing the difference by the standard deviation. P-values were calculated from the negative of the absolute value of the z-scores using the pnorm function in R.^24^ All scripts and data used for the analysis are published on GitLab (https://git.ecdf.ed.ac.uk/twillia2/indirect_effects_covid-19_open_data). This study is reported in accordance with the STROBE statement.^25^

#### Role of the funding source

There was no funding source for this study.

## Results

### Reduction in primary care out of hours consultations and emergency department attendances

The onset of lockdown on the March 23 (occurring one day after the end of epidemiological week 12 of 2020, Figure 1A) until the end of the study period was associated with an almost two-thirds reduction in the number of unscheduled primary care attendances (OR 0·36, 95% CI 0·35-0·37, p < 0·001) when epidemiological weeks 13 to 32 were compared to equivalent dates in 2016-2019 (Figure 2A). For the same period, there was a reduction in emergency department attendances compared to previous years (OR 0·49, 95% CI 0·48-0·49, p < 0·001) (Figure 2B). Comparing OOH consultations to ED attendances, the fall in the latter following lockdown was more marked.

**Figure 2.**
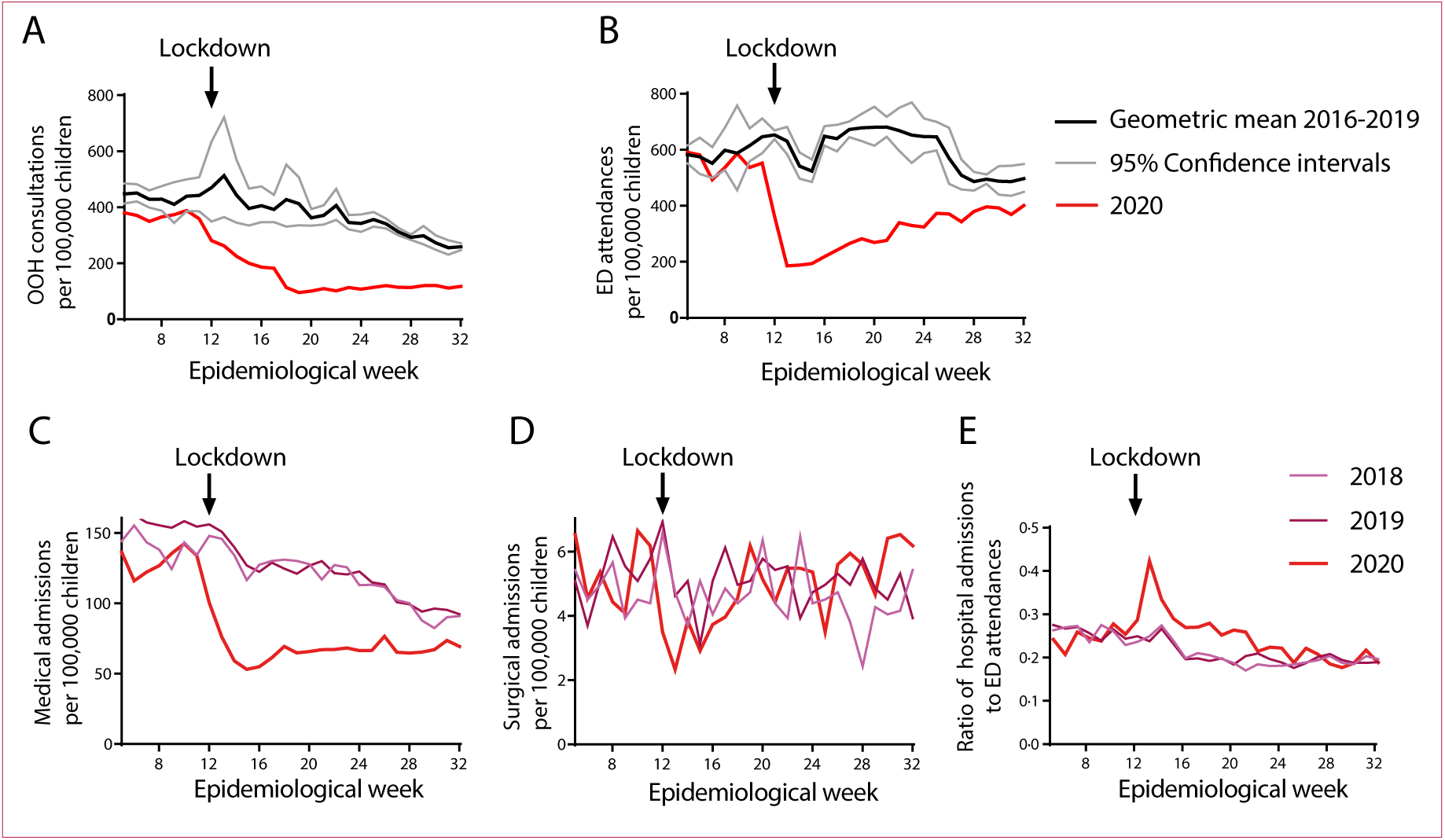
Primary care out of hours consultations and emergency department attendances fell after lockdown. (A) Out of hours (OOH) unscheduled primary care consultations. Number of unscheduled primary care consultations by epidemiological week compared to mean number of consultations, with 95% confidence intervals, for 2016-2019. Data shown for total of 273,455 consultations, including telephone consultations. (B) Emergency department (ED) attendances. Data shown as for (A), for total of 462,437 attendances. (C) Emergency medical paediatric admissions. Number of emergency medical paediatric admisions in 2020, compared to admissions in 2018 and 2019. Data shown for total of 51,596 admissions. (D) Emergency surgical paediatric admissions. Data presented as in (C), for total of 2,480 admissions. All data in A-D normalized by population at risk per year. (E) Proportion of hospital admissions to ED attendances.

### Reduction in emergency hospital admissions for medical but not surgical diagnoses

Comparison of nationwide emergency medical paediatric admissions showed a reduction in these during the lockdown period (Figure 2C), with an odds ratio (OR) of 0·56 for an admission occurring in in epidemiological weeks 13-32, compared to equivalent weeks in 2018-2019 (95% CI 0·55 to 0·57, p < 0.001). There was no equivalent reduction in paediatric surgical admissions in the post-lockdown period overall, with an OR for admission of 1·03 (95% CI 0·95 to 1·12, p= 0·46) (Figure 2D).

### Proportion of Emergency Department attendances to hospital admission during lockdown

As a marker for seriousness of symptoms on presentation to the Emergency Department, the proportion of the number of children admitted as an emergency to the number who attended the emergency department was calculated (for data from 2018-2020) (Figure 2E). In the early weeks of lockdown, there was a rise in the proportion of emergency department attendances compared to numbers admitted to hospital, with a peak in the first week of lockdown (epidemiological week 13, March 23-29). In this week there was a 1·74-fold likelihood of an emergency admission as a proportion of ED attendances, compared to 2018-2019 (95% CI 1·57-1·92, p < 0·001, Fisher’s exact test). Comparing ED activity and the proportion of hospital admissions to ED attendances, by week 22 activity for ED attendances was still at a lower rate than the average for 2016-19 (Figure 2B), but the proportion of admissions to attendances was back to normal levels (Figure 2E).

### Reduction in emergency admissions to PICU for children requiring invasive mechanical ventilation but no increase on severity scores on presentation

The lockdown period beginning March 23, 2020 was associated with a decrease in emergency admissions to PICU requiring invasive mechanical ventilation. Comparison of the number of events in epidemiological weeks 13-26 (March 23 to June 30, 2020; Figure 3A) with equivalent periods in 2016-2019 showed that the OR of an emergency admission requiring invasive mechanical ventilation during the lockdown study period was 0·52 (95% CI 0·37-0·70, p <0·001). The most marked decrease (by 77%) was seen in admissions for disorders of the respiratory system (ICD-11 Chapter 12) (p<0.001) (Figure 3B). There was also a decrease in admissions for disorders of the neurological system (ICD-11 Chapter 8), p=0·01, Supplementary Table 1). No change was seen when compared to previous years in admissions for injury, poisoning or certain other consequences of external causes (ICD-11 Chapter 22, Supplementary Table 2). The PIM2R score for PICU emergency admissions requiring invasive mechanical ventilation during the lockdown study period did not differ from the equivalent epidemiological weeks in 2016-2019 (p-value = 0·23, Kolmogorov-Smirnov test) (Figure 3C). During the lockdown study period there were no emergency admissions requiring invasive mechanical ventilation for bronchiolitis, lower respiratory tract infection or respiratory failure (Table 1).

**Table 1.** Emergency admissions requiring invasive mechanical ventilation falling within ICD-11 Chapter 12 (Disorders of the respiratory system). PICU admissions from March 23 to June 30, 2020, compared to the same date range in previous years (2016-2019).

| **Diagnosis** | **2016** | **2017** | **2018** | **2019** | **2020** |
| --- | --- | --- | --- | --- | --- |
| Bronchiolitis | 10 | 8 | 12 | 9 | 0 |
| LRTI - cause unspecified | 6 | 9 | 9 | 8 | 0 |
| Respiratory Failure - cause unspecified | 4 | 3 | 0 | 0 | 0 |
| Asthma | 0 | 2 | 2 | 0 | 0 |
| Pleural Effusion | 1 | 0 | 0 | 0 | 2 |
| Pneumonia | 1 | 5 | 0 | 4 | 1 |
| Pneumothorax | 0 | 0 | 0 | 0 | 1 |
| Pulmonary Oedema | 0 | 1 | 0 | 1 | 1 |
| Tracheitis | 0 | 0 | 0 | 0 | 1 |
| Other | 5 | 9 | 5 | 11 | 1 |

**Figure 3:**
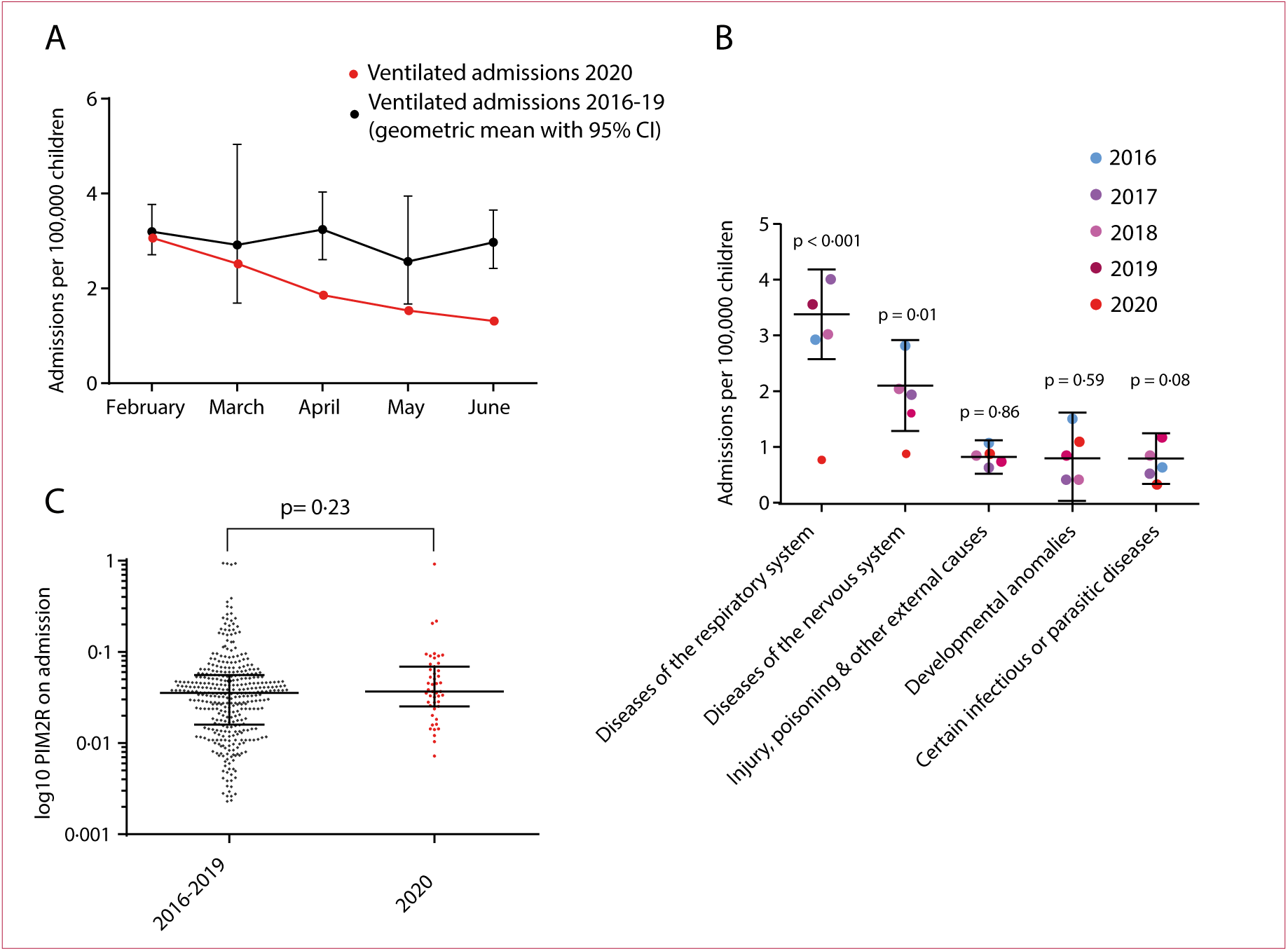
PICU admissions. (A) The overall number of emergency admissions to PICU requiring invasive mechanical ventilation in March-June 2020 fell when compared to the mean for previous years. Geometric mean and 95% CI shown for 2016-2019, expressed as number of admissions per 100,000 children at risk. Data shown for total of 413 admissions during the study period. (B) The reduction in admissions was most marked for diseases of the respiratory system. Geometric mean and 95% CIs shown for five most common ICD-11 chapters. There was a reduction in overall admissions for respiratory causes (77% reduction), and no change in admissions for injury, poisoning or other external causes. P-values calculated from z-scores for 2020 compared to means and standard deviation for 2016-2020. (C) A reduction in admissions was not associated with an increase in severity scores on arrival in the PICU. Median and interquartile ranges shown for log_10_ PIM2R scores on admission for admissions between March 23 and June 30, 2020 compared with the equivalent period in 2016-2029. Distributions compared using a Kolmogorov-Smirnov test, p = 0·23.

### Childhood mortality

Childhood mortality in epidemiological weeks 13-30 in 2020 did not differ significantly from mean in equivalent weeks in 2016-19, for overall deaths, deaths in ages 0-4 years, and deaths in ages 5-14 years (Figure 4A). Of note, the cause of death for a proportion of cases in years 2019 (10/70) and 2020 (14/78, Supplementary Table 3) had not yet been finalised and therefore could not be included (Figure 4B, grey bars).

**Figure 4:**
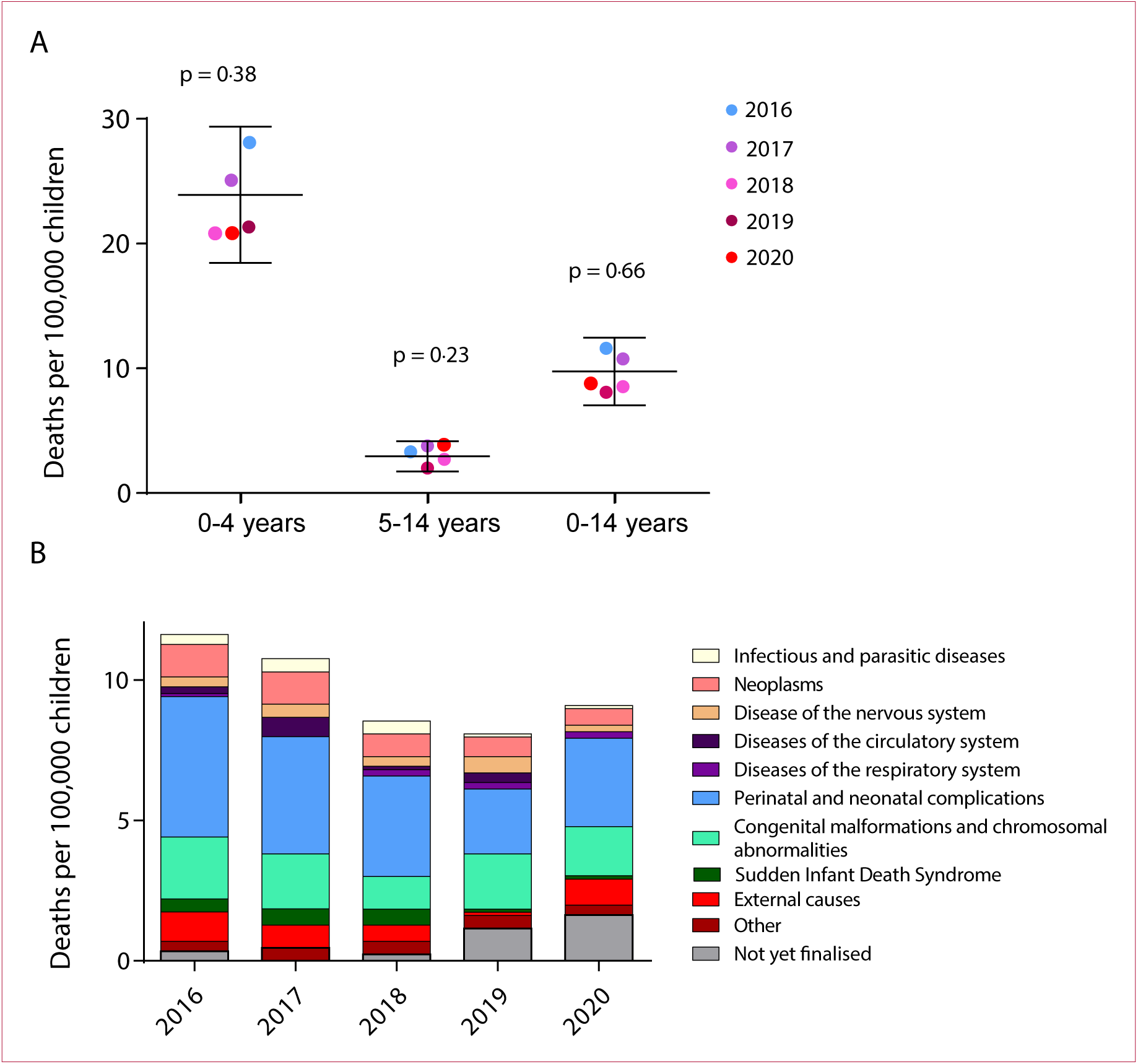
Childhood mortality during lockdown study period. (A) There was no significant change in child mortality in the lockdown study period compared to the equivalent period in previous years. Number of deaths per 100,000 children in Scotland in epidemiological weeks 13-30 for 2020, compared to the 2016-2019. Horizontal lines show means with 95% CIs for years 2016-2019. P-values calculated from z-scores for 2020 compared to means and standard deviation for 2016-2020. (B) Childhood deaths by cause in 2020 compared to previous years. Deaths per 100,000 children classified by ICD-10 chapter. A number of deaths for 2019 and 2020 have not yet been classified by ICD-10 code and fall into the “Not yet classified” group.

## Discussion

Overall emergency paediatric healthcare utilisation by children in Scotland reduced significantly during the lockdown period with no immediate, measurable severe adverse effects on physical child health at a population level observed. Specifically, a reduction in out-of-hours primary care consultations, ED attendances and unplanned hospital admissions did not result in increased disease severity on presentation to PICU or mortality.

A significant reduction was seen in the number of PICU admissions for respiratory disease and those requiring invasive mechanical ventilation during the lockdown study period, supporting recent reports,^11^ consistent with NPIs being associated with an overall reduction in non-COVID-19 related respiratory infections. There were no PICU admissions for bronchiolitis, and a dramatic reduction in other lower respiratory tract infections across Scotland during this time. During the same period, no increase was seen in children admitted to PICU within the category of injury, poisoning, or other external causes”, which may have been expected if lockdown was associated with reduced surveillance by health, education and social care workers. No significant changes were seen in paediatric mortality for this period across any age group examined.

The causes for the observed reduction in rates of paediatric emergency healthcare utilisation during the lockdown study period are likely to be multifactorial. Changes in patterns of interaction between children, shielding measures for at risk groups, and restricted domestic and international travel during the lockdown study period are likely to have reduced the transmission rates of infectious pathogens, and may have reduced exposure to some of the triggers for respiratory diseases (e.g, air pollution in asthma).^10^ Similarly, increased parental supervision may have improved adherence with medical therapies and thus reduced exacerbation frequency by improved compliance with prescribed medications, such as anticonvulsants. A significant reduction in admissions to PICU for neurological disorders was seen (Figure 3B), alongside a reduction in the number of diagnoses of seizure (Supplementary Table 1).

The observed changes in health care utilisation outlined in this study appear mainly related to medical issues since surgical presentations remain relatively unchanged (Figure 2C/D).This discrepancy between medical and surgical presentation rates suggests that the interruption of pathogen transmission by NPIs was the most significant factor in reducing paediatric medical admissions rather than an alteration in parental care seeking behaviour during the lockdown period. In addition, in first weeks of lockdown, a higher proportion of admissions to hospital compared to emergency department attendances was observed (Figure 2E). One explanation for this is that whilst healthcare-seeking behaviour changed extremely quickly, the impact of NPIs on pathogen circulation (and thus on presentations with infectious diseases) was slower, resulting in fewer presentations with the same amount of underlying hospitalisable morbidity, thus increasing relative admission rates at this specific point in time.

Despite initial concerns in the literature regarding delayed presentations for non-SARS-CoV-2 related conditions, for example diabetic keto-acidosis and sepsis,^28^ and delayed new cancer diagnoses,^13^ our study shows that these changes have not translated into a measurable increase in severe paediatric morbidity or mortality in Scotland. Overall, the data supports a reduction in the burden of childhood disease across Scotland. However, there is growing body of evidence that the lockdown period may have a detrimental effect, both in the short- and long-term, particularly on the mental health of children,^29^ a topic that is beyond the scope of this current study.

Our study uses high quality, standardised population data from a variety of sources which address the study question from a number of different viewpoints. The use of complete national datasets allowed us to examine the entire paediatric population of Scotland during the lockdown study period and compare this with data collected in previous years. The results from all analyses are concordant with each other, highlighting a picture of reduced healthcare seeking without short-term increase in severe paediatric disease or mortality. To our knowledge, this is the first national study to quantify the indirect effects of the COVID-19 pandemic on severe paediatric morbidity and mortality.

Whilst the analysis presented is generally reassuring, we acknowledge that it represents a broad view of child health in Scotland and population level data may fail to represent individual patient experiences. However, any delayed presentations to hospital with more severe symptoms, did not appear to result in increased risk of PICU admission or death. Minor differences in the coding of PICU admission (ICD-11) and causes of mortality (ICD-10) may make these groups harder to compare. Similarly, mortality data was incomplete for 2019-20, with some deaths still to be attributed a cause. Thus, it is possible that these missing deaths (Supplementary Table 3), once coded, could influence the findings of our analyses. In addition, our analysis only covers the acute lockdown period and therefore the longer term impact on physical health, for example the effects of cancellation of routine surgery or outpatient clinics, cannot be quantified. The fundamental changes made to primary care provision (eg. fewer face to face GP consultations and use of virtual patient assessments) as well as the effect of Government/public health messaging on healthcare-seeking behaviour are also important factors but out-with the scope of this report. We must also be cognisant that physical health is by no means the only parameter of child health. In particular, our study does not address the impact on loss of learning opportunities, mental health or the impact of lockdown on the safety of vulnerable children, none of which should be underestimated.

National lockdown did not appear to result in measurable harm at the most severe end of the disease spectrum (PICU admission and death). However, follow-up of the direct long term effects of paediatric infection with SARS-CoV-2, and the impact of physical distancing measures on wider indicators of physical and mental health, including missing periods of education, must be addressed to ensure that health policy takes account of holistic impacts of the SARS-CoV-2 pandemic on paediatric health. However, a deeper understanding of this period offers the opportunity to consider how to safely re-structure health care services provision. Discussions in this area will be supported by further studies delineating what proportion of the changes in care seeking related to reduction in disease burden, or to changes in care seeking behavior. Going forwards, patients, caregivers, and health care providers may seek to continue novel ways of managing both acute and chronic disease, including optimisation of virtual consulting and point of care home measurements.

## Conclusion

Our analysis shows a reduction in paediatric emergency care-seeking utilisation that occurred as a consequence of the widespread adoption of NPIs and societal lockdown measures. This reduction is likely to be due to a combination of changes in health care seeking behavior, and a fall in overall burden of infectious causes of childhood disease. These measures do not appear to have been associated with evidence of severe harm to children in Scotland during the lockdown period.

## Data Availability

The source code for all the analyses, alongside the anonymized datasets used in these, has been published on GitLab (https://git.ecdf.ed.ac.uk/twillia2/indirect_effects_covid-19_open_data).

## Acknowledgments

We would like to thank Julie Ramsay and Marie Kay from National Records of Scotland, and Kathy McGregor and Jaime Villacampa from Public Health Scotland, for their help in securing the data underlying these analyses, and for their comments on the manuscript.

## ICMJE Statement

RJL and TCW conceived the study. CM, OS, SC, PD, NG, ST, DU and BG made substantial contributions to the design of the work. CL, RL, CM, JM, IP, AT and MVD contributed towards acquisition of data for the work. TCW, CM and HH performed the analysis and interpretation of the data. RJL, TCW, CM and OS drafted the manuscript; SC, PD, NG, ST, DU, BG, CL, RL, CM, JM, IP, AT and MVD revised it critically for important intellectual content. All authors gave final approval of the version to be published and agree to be accountable for all aspects of the work in ensuring that questions related to the accuracy or integrity of any part of the work are appropriately investigated and resolved.

## Declaration of interests

The authors have no conflict of interests to declare.

## Funding

TCW is the recipient of a Wellcome Trust Award [204802/Z/16/Z].

## Supplementary Tables

**Supplementary Table 1.** Emergency paediatric intensive care unit (PICU) admissions requiring invasive mechanical ventilation, diagnoses within ICD-11 Chapter 18 (disorders of the neurological system). PICU admissions from March 23 to June 30 2020, compared to the same date range in previous years (2016-2019).

| **Diagnosis** | **2016** | **2017** | **2018** | **2019** | **2020** |
| --- | --- | --- | --- | --- | --- |
| Seizure disorder | 22 | 14 | 15 | 12 | 8 |
| Autoimmune | 1 | 0 | 0 | 0 | 0 |
| Infection | 0 | 1 | 0 | 0 | 0 |
| Inflammatory | 1 | 0 | 0 | 0 | 0 |
| Intracranial Haemorrhage /<br>Ischaemia | 0 | 0 | 1 | 1 | 0 |
| Headache | 0 | 0 | 1 | 0 | 0 |
| Hydrocephalus | 1 | 0 | 0 | 0 | 0 |
| Neuropathy | 0 | 0 | 0 | 1 | 0 |
| Motor Neurone Disease | 0 | 0 | 1 | 0 | 0 |
| Intracranial Haemorrhage / Toxic | 0 | 1 | 0 | 0 | 0 |
| Other | 1 | 1 | 1 | 1 | 0 |

**Supplementary Table 2.** Emergency paediatric intensive care unit (PICU) admissions requiring invasive mechanical ventilation, diagnoses within ICD-11 Chapter 22 (injury, poisoning or certain other consequences of external causes). PICU admissions from March 23 to June 30, 2020, compared to the same date range in previous years (2016-2019).

| **Diagnosis** | **2016** | **2017** | **2018** | **2019** | **2020** |
| --- | --- | --- | --- | --- | --- |
| Trauma - Head | 4 | 4 | 3 | 3 | 4 |
| Trauma - Intra-abdominal | 0 | 1 | 1 | 0 | 1 |
| Trauma - Multiple Injuries | 2 | 0 | 3 | 1 | 0 |
| Trauma - Thoracic | 2 | 0 | 0 | 0 | 0 |
| Toxic | 0 | 1 | 0 | 1 | 1 |
| Other | 2 | 0 | 1 | 2 | 2 |

**Supplementary Table 3.** Deaths of under 15 year olds registered in Scotland during epidemiological weeks 13 to 30 in 2016, 2017, 2018, 2019 and 2020. Including the chapter (ICD-10) containing the underlying cause of death and age groups.

| **Diagnosis** | **2016** | **2017** | **2018** | **2019** | **2020** |
| --- | --- | --- | --- | --- | --- |
| Infectious and parasitic diseases | 3 | 4 | 4 | 1 | 1 |
| Neoplasms | 10 | 10 | 7 | 6 | 5 |
| Disease of the nervous system | 3 | 4 | 3 | 5 | 2 |
| Diseases of the circulatory system | 2 | 6 | 1 | 3 | 0 |
| Diseases of the respiratory system | 1 | 0 | 2 | 2 | 2 |
| Perinatal and neonatal complications | 43 | 36 | 31 | 20 | 27 |
| Congenital malformations and chromosomal abnormalities | 19 | 17 | 10 | 17 | 15 |
| Sudden Infant Death Syndrome | 4 | 5 | 5 | 1 | 1 |
| External causes | 9 | 7 | 5 | 1 | 8 |
| Other | 3 | 4 | 4 | 4 | 3 |
| Not yet classified | 3 | 0 | 2 | 10 | 14 |
| **Total** | **100** | **93** | **74** | **70** | **78** |

## Notes

### Competing Interest Statement

The authors have declared no competing interest.

### Author Declarations

Disclosure controlled, aggregate count data of relevant healthcare utilisation events (apart from PICU admissions) was provided by Public Health Scotland (PHS). The data used is publicly available upon request, with no ethical permissions required. Analysis of the PICU data was performed as part of a service evaluation program and in concordance with both NHS Greater Glasgow and Clyde and NHS Lothian Quality Improvement policies ethical approval is not required.

